# Overcome the Limitation of Phenome-Wide Association Studies (PheWAS): Extension of PheWAS to Efficient and Robust Large-Scale ICD Codes Analysis

**DOI:** 10.1101/2024.04.15.24305098

**Authors:** Ya−Chen Lin, Siwei Zhang, Tess Vessels, Lisa Bastarache, Cosmin Adrian Bejan, Ryan S Hsie, Elizabeth J Philips, Doug M Ruderfer, Jill M. Pulley, Todd L Edwards, Quinn S Wells, Jeremy L Warner, Joshua C Denny, Dan M Roden, Hakmook Kang, Yaomin Xu

## Abstract

The Phenome-wide association studies (PheWAS) have become widely used for efficient, high-throughput evaluation of relationship between a genetic factor and a large number of disease phenotypes, typically extracted from a DNA biobank linked with electronic medical records (EMR). Phecodes, billing code-derived disease case-control status, are usually used as outcome variables in PheWAS and logistic regression has been the standard choice of analysis method. Since the clinical diagnoses in EMR are often inaccurate with errors which can lead to biases in the odds ratio estimates, much effort has been put to accurately define the cases and controls to ensure an accurate analysis. Specifically in order to correctly classify controls in the population, an exclusion criteria list for each Phecode was manually compiled to obtain unbiased odds ratios. However, the accuracy of the list cannot be guaranteed without extensive data curation process. The costly curation process limits the efficiency of large-scale analyses that take full advantage of all structured phenotypic information available in EMR. Here, we proposed to estimate relative risks (RR) instead. We first demonstrated the desired nature of RR that overcomes the inaccuracy in the controls via theoretical formula. With simulation and real data application, we further confirmed that RR is unbiased without compiling exclusion criteria lists. With RR as estimates, we are able to efficiently extend PheWAS to a larger-scale, phenome construction agnostic analysis of phenotypes, using ICD 9/10 codes, which preserve much more disease-related clinical information than Phecodes.

## 1 INTRODUCTION

The first successful Genome-Wide Association Study (GWAS) was published by Ozaki et al. [1]. Since then, GWAS has been used as a main tool to identify new genetic association in many diseases. With the accumulation of DNA biobank linked with electronic medical records (EMR), large number of genetic-disease association can be conducted in one study. One alternative approach to assess the genetic-disease association is the Phenome-Wide Association Study (PheWAS), proposed by Denny et al. [2] and has demonstrated reproducibility from known association on GWAS catalog [3]. Following the concept of “reverse-GWAS”, PheWAS analyses scan through large number phenotypes with given genetic variant.

Current practice for PheWAS analyses models the genetic-disease association with logistic regression consisting binary disease outcomes and genetic variant exposure of patients. In the EMR system, ICD codes remain the most commonly used phenotype outcomes to assess patient’s disease status. According to World Health Organization (WHO), ICD 9 codes have been used from 1900 till now. Currently there are around 13000 codes available. Starting October 2015, ICD10 codes have officially entered the health system. The new ICD10 system will carry around 68000 codes with more classification options to categorize diseases. However, the billing codes are known to be noisy. One assumption for logistic regression is to have clear definition of case and control groups. Since 2010, several studies have been done with raw ICD 9 codes or combined ICD 9 codes [4][5]. Although showing some degree of reproducibility, violating the logistic regression might result in biased results. In 2016, Denny, Bastarache, and Roden et al. [6] proposed to group ICD 9 codes into “Phecodes” based on the similarities of the different ICD 9 codes. To be classified as a “case” for a Phecode, the subject needs to have at least 2 or more ICD 9 codes on different days mapping to that specific Phecode. Since most of the efforts in clinical practice have been spent on trying to define true cases, the “control” population might not be pure controls. Therefore, one exclusion criteria list was composed manually for each Phecode, considering all possible related conditions to the Phecode to try to filter out the true controls. To be classified as a “control” for a Phecode, the subject cannot have any ICD 9 code mapping to the Phecode and not have any ICD 9 code in the exclusion criteria list for the Phecode.

From the clinical standpoint, manually compiled exclusion criteria list, considering disease comorbidity, might perform better when filtering out true controls, acting as Gold-standard. However, there are few drawbacks with this approach. First, the disease network is large and complex and still, there are unknown relationships between the diseases. It is highly possible that the complete disease network wasn’t considered fully. Secondly, the time required to manually compile these exclusion criteria lists might be long. Although it might be true that the grouped ICD codes could give a better understanding of the general view of the disease structure, grouping results in loss of information. The ICD codes provide more details on the disease status, treatment information and so on. With the time-consuming aspect of the Gold-standard lists, it will be difficult to generalize this method to larger number of codes, common to large public databases like UK biobank.

To overcome the limitation, we turned to the original study design. In case control studies, the data are collected based on the disease status. In contrast, retrospective studies are accessed after some patients have already developed the outcomes. The investigators then jump back in time to identify the exposure status at a point of time before any development of the outcome. Then, one can determine whether the subject subsequently develop the outcome. In PheWAS, we have one exposure, usually the SNPs, with multiple disease outcomes. The proportion of subjects that are in exposed and unexposed groups are the same for each SNP-outcome analysis. Further, SNPs exist before the development of the general diseases in the system. Thus, we can think of population divided by the exposure status and followed throughout time in the EMR system. This kind of study can be also viewed as a retrospective cohort study. Once we established the study design, we can further explore other options to assess the genetic-disease association. In case control studies, since the investigators already set up the disease prevalence, odds ratio remains the only measurement. In contrast, in retrospective cohort studies, relative risk is also a valid measurement. In this study, we evaluated the usage of relative risk as the measurement in PheWAS analyses and demonstrated that it can overcome the control misclassification issue and bypassed the exclusion criteria limitation.

In Theoretical Background section, we showed the theory of how relative risk can be free of biases resulting from the misclassification. In Simulation section, we showed the performance of relative risk comparing to Gold-standard odds ratios with several combination of outcome prevalence and degrees of misclassification. In Real Data Analysis section, we illustrated with real data that relative risk model behaved similarly to Gold-standard model. Further, we generalized PheWAS to ICD 9 codes. We were able to obtain further useful disease information that was missed in Phecodes analysis.

## 2 THEORETICAL BACKGROUNDS

To begin with, we would like to explore different scenarios with theoretical formula derivation. Table 1 illustrates the distribution of hypothetical data with binary exposure (E) and binary outcome (Y). Letter a, b, c and d denote the number of observations in each E, Y combination. The sum of a, b, c, d equals the number of total observations (N).

**Table 1:**
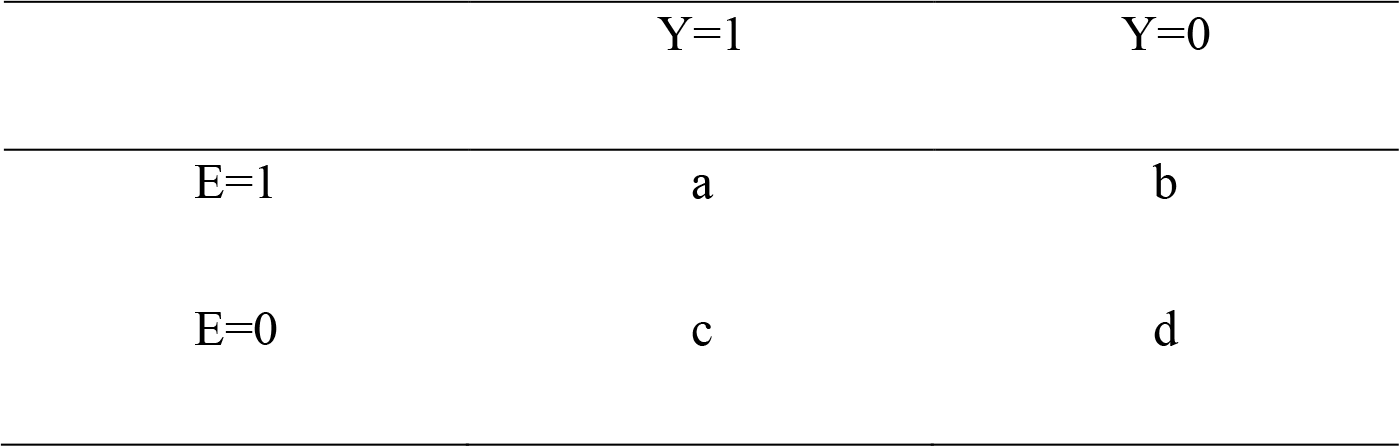
2 × 2 table with Outcome and Exposure in hypothetical data.

With table 1, the formula for odds ratio is 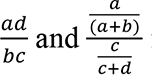 for relative risk. The odds ratio can be converted to relative risk with formula (Grant [7]) 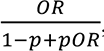, where p denotes the risk in the control groups, in this case,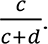. There are two scenarios that could happen when defining cases and controls: cases misclassified into controls and controls misclassified into cases. For the purpose of PheWAS analyses, the scenario where cases are misclassified into controls is the primary focus and shown in the main text. The method and simulation result for the scenario where controls are misclassified into cases is shown in Table2.

We can derive the formula when cases were misclassified into controls with Table 2. We denote Z as the proportion of cases being misclassified as controls and T as the proportion of the misclassification NOT being removed.

**Table 2:** 2 × 2 table with Outcome and Exposure; cases misclassified as controls.

|  | Y=1 | Y=0 |
| --- | --- | --- |
| E=1 | a - aZ | b + aTZ |
| E=0 | c - cZ | d + cTZ |

The formula for odds ratio becomes 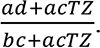 When misclassification is completely removed (T=0), odds ratio is 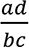, unbiased and free of Z. When the estimated unbiased odds ratio is converted to relative risk, the estimated relative risk is 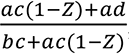, a function of Z.

When misclassification is completely ignored (T=1), odds ratio is 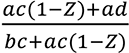, biased and a function of Z. The bias increases as Z increases. It’s worth noting that when Z approaches 1, the biased odds ratio approaches 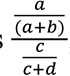, the relative risk. When the estimated biased odds ratio is converted to relative risk, the estimated relative risk is 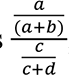, unbiased and free of Z. For the direct estimation of relative risk, with the goal of our paper, we do not remove any observation (T=1). The estimated relative risk is 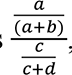, unbiased and free of Z.

With the theoretical formula, the estimated odds ratios are unbiased only when all misclassification is removed (T=0). The direct estimation of relative risk or conversion from biased odds ratios where no observation was removed in the original dataset (T=1) produce unbiased relative risk.

## 3 SIMULATION

### 3.1 Univariate Simulation

To validate the theoretical formula, we first conducted univariate simulation. In univariate simulation, we included only a binary exposure (E) as a predictor and a binary outcome (Y). We denote proportion of cases being misclassified as controls as Z. The probability of exposure (Pe) was set to 0.1. The prevalence of Y (P1) includes 0.02 and 0.3 to explore the influence of the prevalence on the estimates. The true relative risks (RR) were set to 0.5 and 3. Four P1 and RR combinations are listed as below:

To explore the influence of varying levels of misclassification (Z), in each P1 and RR combination, 5 conditions of Z were implemented, including 0 (No misclassification), 0.1, 0.25, 0.5 and 0.75 (75% of the true cases misclassified into controls). 500 simulations and 7000 observations (N) were conducted for each P1 and RR combination.

The distribution for binary E follows Binomial (N, 0.1). The conditional probabilities, *P*(*Y* = 1|*E* = 1) and *P*(*Y* = 1|*E* = 0) were calculated with pre-specified *Pe*, *P*1 and *RR* according to the above list. With Bayes rules, the conditional probabilities can be computed with known joint probabilities with the relative risk formula where *P* (*Y* = 1, *E* = 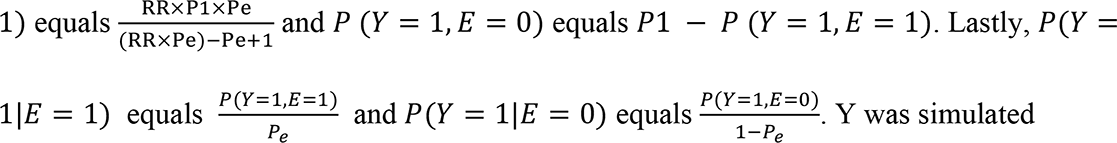 according to the conditional probabilities, *P*(*Y* = 1|*E* = 1) and *P*(*Y* = 1|*E* = 0). By directly calculating the conditional probabilities, we can easily make sure all the 4 conditional probabilities, *P*(*Y* = 1|*E* = 1), *P*(*Y* = 1|*E* = 0), *P*(*Y* = 0|*E* = 1) and *P*(*Y* = 0|*E* = 0), are positive. After E and Y were simulated, Z proportion of cases (Y=1) was randomly converted to controls (Y=0). For the additional simulation where controls are misclassified as cases in Supplementary Figure 1, the simulation procedures follow the above univariate simulation with P1 = 0.3 and RR = 0.5. After E and Y were simulated, Z proportion of controls (Y=0) was randomly converted to cases (Y=1).

### 3.2 Multivariate Simulation

We included a multivariable simulation mimicking the commonly used additive modeling of SNPs for the exposure variable and the presence of a continuous covariate (C). The data were simulated with a log-binomial model: *log* (*P*(*Y* = 1|*E*, *C*)) = *log*(0.15) + *log*(2)*E* + *log*(1.3)*C*. The exposure variable E was simulated with categories 0, 1, 2 indicating homozygous dominance, heterozygotes, and homozygous recessive genotypes. The corresponding probabilities were 0.6, 0.3 and 0.1. The covariate C was simulated with Normal distribution, mean = 0 and variance = 0.3. The prevalence of Y was approximately 0.456, empirically. Similar to univariate simulation, Z proportion of cases (Y=1) was randomly converted to controls (Y=0).

### 3.3 Statistical Models

For each simulation, we included three models for relative risk comparison, bias-corrected (Firth [8]) Poisson, Logistic regression with exclusion criteria and Logistic regression without exclusion criteria. For the logistic regression models, the odds ratios were converted to relative risk with the conversion formula presented in Theoretical Background section. We also reported comparison of Logistic regression with exclusion criteria and Logistic regression without exclusion criteria for odds ratios. Logistic regression with exclusion criteria is the current practice of PheWAS, also referred to as the Gold-standard model. Logistic regression was conducted on dataset that the misclassification is manually removed. In our simulation, we assumed that the Gold-standard model can remove all misclassification (T=0). Logistic regression without exclusion criteria refers to conducting logistic regression on dataset without removing any observation (T=1). For Poisson model, the regression was also conducted on the dataset without removing any observation.

### 3.4 Simulation Results

Following the formula in Theoretical Background section, when cases are misclassified into controls, odds ratio from Logistic regression with exclusion criteria stays unbiased with varying degree of misclassification. When misclassification is not completely removed, odds ratio is biased and a function of Z. As misclassification rate increases, biased odds ratio approaches the true relative risk. Therefore, if the true odds ratio and true relative risk differ more, the biases should become more profound.

The univariate simulation results are shown in Figure 1 and Figure 2 with boxplots comparing the proportion of cases misclassified as controls (X axis) and the estimates for odds ratios/relative risk (Y axis). As shown in Figure 1, when the prevalence is 0.02, the difference between the true relative risk and the true odds ratio is small (0.5 vs. 0.495 and 3 vs. 3.105). As Z increases, biases in odds ratios are almost negligible when ignoring the misclassification. However, when the prevalence becomes 0.3, the true relative risk and odds ratio differ more (0.5 vs. 0.406 and 3 vs. 9), the biases become noticeable as Z increases. The biased odds ratios bias toward the true relative risk when ignoring the misclassification. From Figure 1, unbiased odds ratios can only be obtained with logistic regression when exclusion criteria were applied (all misclassification is removed).

**Figure 1:**
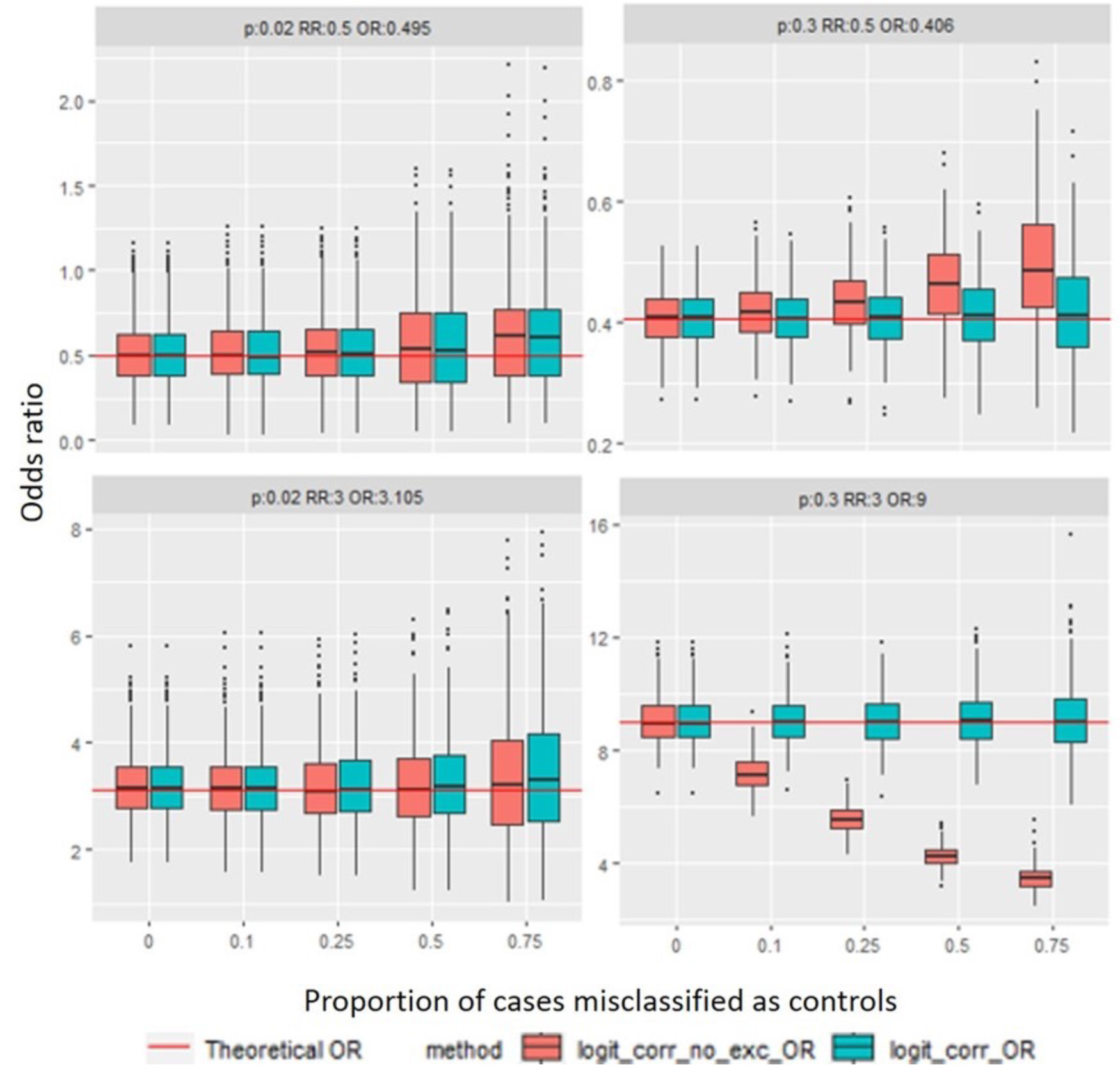
Univariate simulation result: Odds ratio boxplots for prevalence = 0.02, 0.3 and exposure probability = 0.1 with varying true odds ratios. The true odds ratios are 0.495 (top left), 0.406 (top right), 3.105 (bottom left) and 9 (bottom right). The x-axis denotes the proportion of cases being misclassified as controls. The y-axis denotes the odds ratio estimates from the logistic regression with (teal boxes) and without exclusion criteria (red boxes).

**Figure 2:**
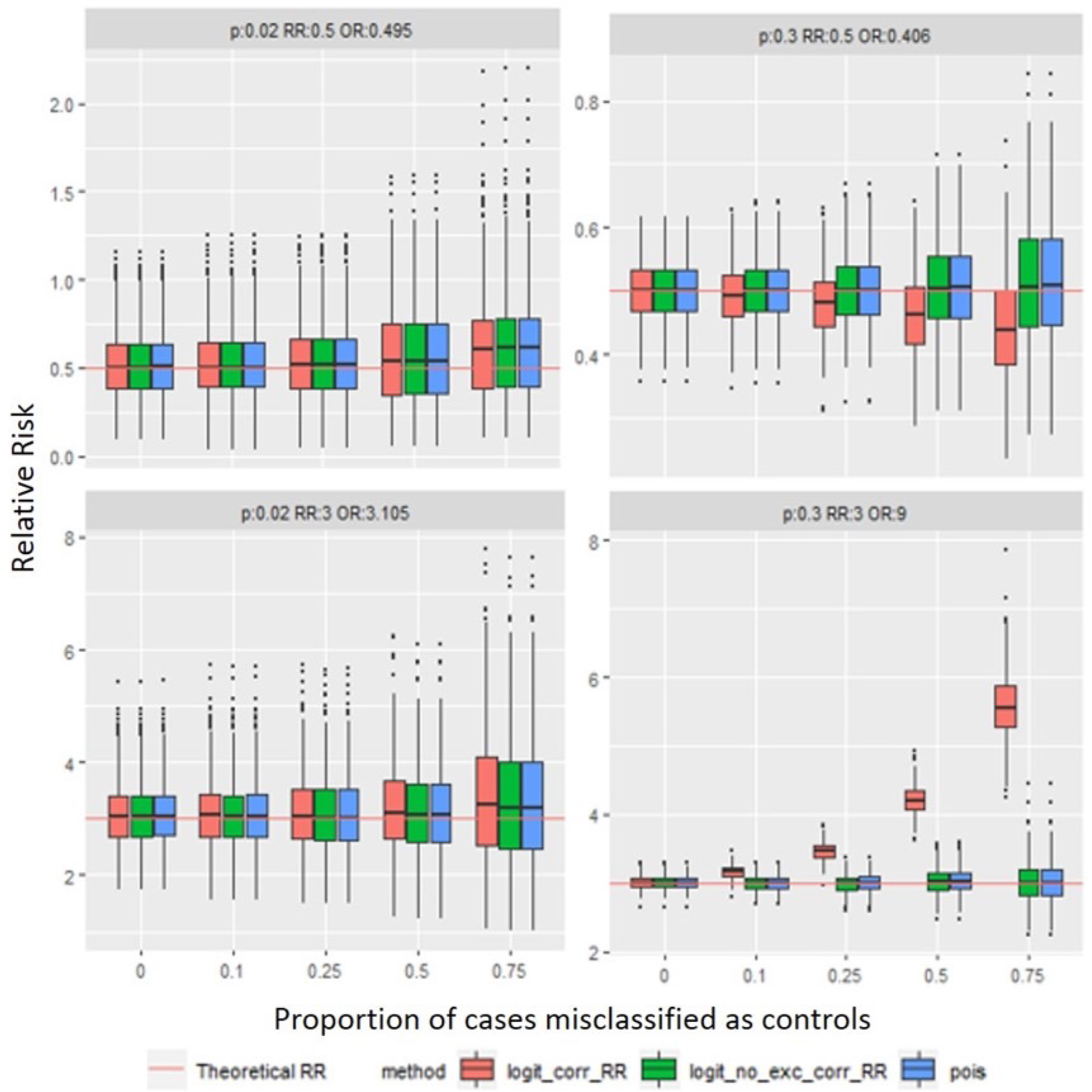
Univariate result: Relative risk boxplots for prevalence = 0.02, 0.3 and exposure probability = 0.1 with varying true relative risk ratios. The relative risk ratios are 0.5 (top row) and 3 (bottom row). The x-axis denotes the proportion of cases being misclassified as controls. The y-axis denotes the relative risk estimates from Poisson model (blue boxes), converted relative risk estimates from logistic regression with (red boxes) and without exclusion criteria (green boxes).

Counter-intuitively, when the estimated unbiased odds ratios were converted to relative risk, the estimates were biased. The magnitudes of biases were shown as a function of Z in Theoretical Background section. The corresponding simulation result is shown in Figure 2. Relative risk converted from unbiased odds ratio from Logistic regression with exclusion criteria is biased. Similar to Figure 1, the biases are not noticeable when the prevalence of Y is smaller. As the prevalence increases to 0.3, the bias becomes profound as Z increases. The relative risk estimated from Poisson model and converted from biased odds ratio stay unbiased in all scenarios. The univariate simulation result confirms the conclusion obtained in Theoretical Background section.

The multivariable simulation result is shown in Figure 3. With the additive modeling of exposure and the presence of covariate C, the odds ratios (Figure 3, top) are unbiased only when all misclassification is removed (T=0), same as what we observe in univariate simulation. In the bottom plot, the estimated relative risk converted from unbiased odds ratio is biased and increases as Z increases. Unbiased relative risk is only obtained in the Poisson model. Interestingly, the relative risk converted from biased odds ratio is biased, unlike what we observe in Figure 2. We hypothesize that the reason for the biases might be due to the added covariate C. The conversion formula presented in Theoretical Background section does not consider the presence of any additional covariate.

**Figure 3:**
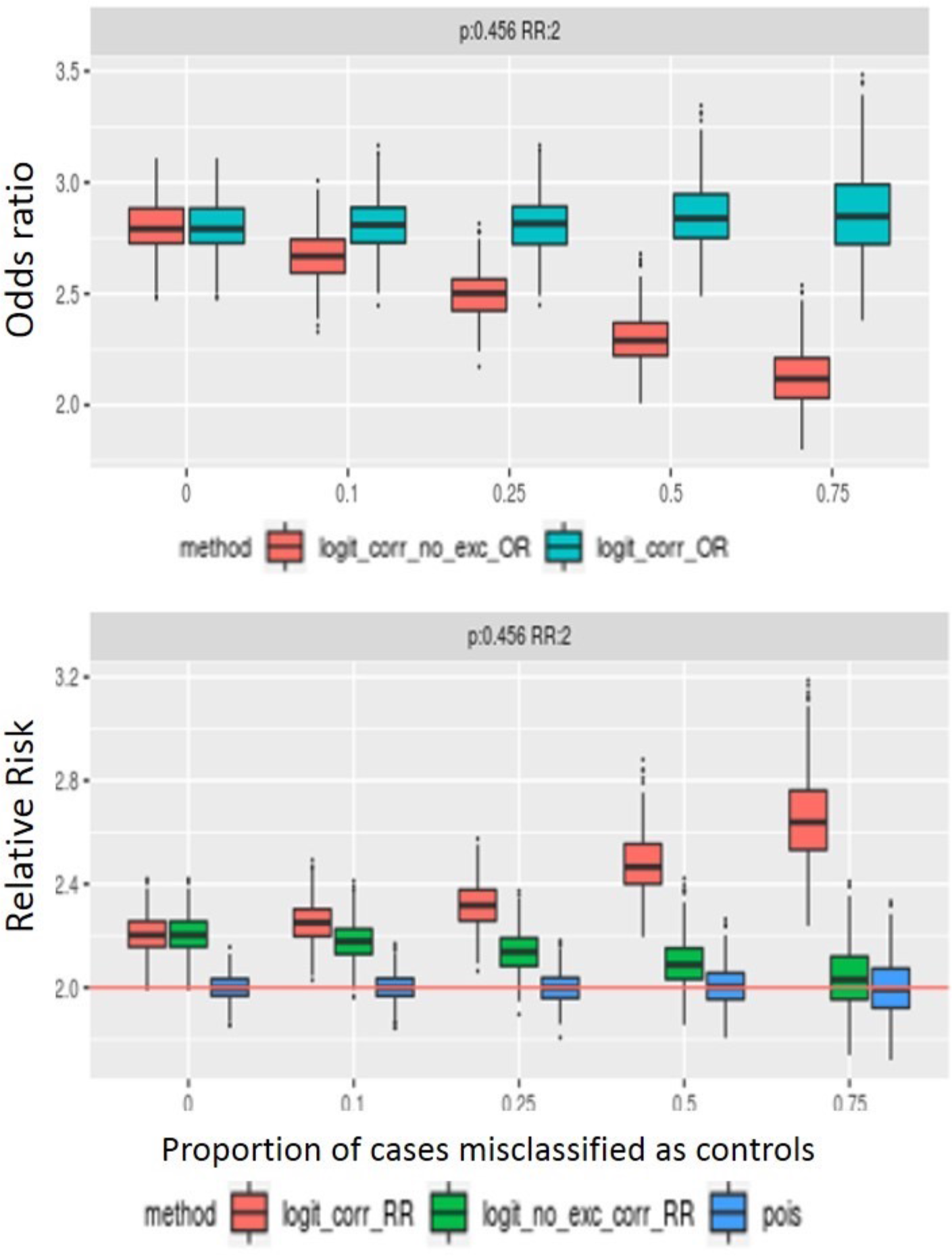
Multivariable simulation result: Odds ratios boxplot (top) and relative risk boxplot (bottom) for exposure probability = 0.1, prevalence = 0.456 and true relative risk = 2. The x-axis denotes the proportion of cases being misclassified as controls. The y- axis for the top figure denotes the odds ratio estimates from the logistic regression with (teal boxes) and without exclusion criteria (red boxes). The y-axis for the bottom plot denotes the relative risk estimates from Poisson model (blue boxes), converted relative risk estimates from logistic regression with (red boxes) and without exclusion criteria (green boxes).

Although not directly pertaining to PheWAS analyses, the scenario where controls were misclassified as cases is shown in Supplementary Figure 1. In Table2, the formula for both relative risk and odds ratio obtained ignoring misclassification are biased and functions of Z. In odds ratio estimation (Supplementary Figure 1, left), exclusion criteria are needed to obtain unbiased odds ratios. In relative risk estimation (Supplementary Figure 1, right), all methods failed to obtain unbiased relative risk.

## 4 CASE STUDY

### 4.1 Data Source

This project was approved by the Vanderbilt University Medical Center (VUMC) Institutional Review Board. The data were obtained from BioVU, the VUMC biobank containing DNA extracted from leftover and otherwise discarded clinical blood specimens selected at random from consenting patients seen within the VUMC healthcare system.

BioVU DNA samples are linked to the Synthetic Derivative (SD), a de-identified research EHR with records spanning from 1990-present, which allows for dense, longitudinal clinical annotation of this genetic data. The SNPs were selected from the previous published paper (Denny et al. [2]) that pass the quality control after imputation. For PheWAS analyses, there are 44764 subjects available from the BioVU cohort. For the Phecodes, following the procedure in Denny et al. [2], we excluded Phecodes with case number less than 20. 1446 Phecodes are available for analyses. The exclusion criteria will be implemented for Gold-standard logistic regression following the published criteria in Denny et al. [2]. For ICD 9 analysis, there are 44846 subjects available from the BioVU cohort. For the ICD 9 codes, we excluded ICD 9 codes with case number < 20. 6329 ICD 9 codes are available for analyses.

### 4.2 Statistical Analysis

To illustrate the potential biases in BioVU data, Logistic regression with and without exclusion criteria were performed with SNP rs14483486 in Figure 4 following a table demonstrating when the biases are the most obvious. The percent biases were calculated with the odds ratios estimated from the Logistic regression model with and without exclusion criteria (100×(OR_without - OR_with)/OR_with). The prevalence and misclassification rate were calculated assuming the observations removed after exclusion criteria were true cases. An example calculation of the prevalence and misclassification rate is given in Clinical Application Results section.

**Figure 4:**
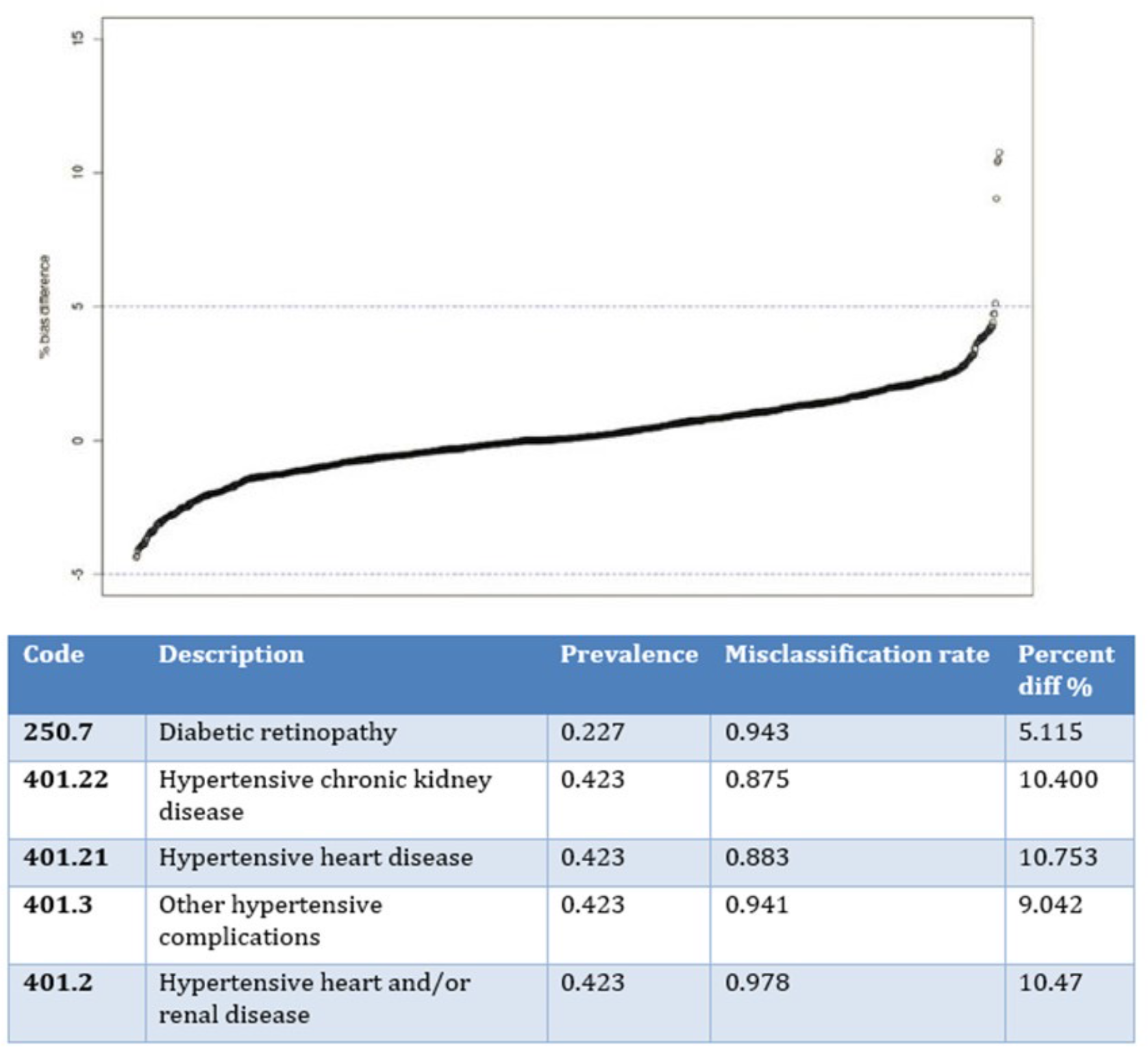
Bias illustration with SNP rs14483486. In the top figure, the y-axis denotes the percent difference of the odds ratios estimated from univariate logistic regression with and without exclusion criteria. We denote this as percent bias as the odds ratios from the Gold-standard model acts as the truth. The percent bias was calculated with formula: (100×(OR_without - OR_with)/OR_with). Each dot represents a Phecode. The bottom table summarizes information, including the Phecode ID, description of the Phecode, prevalence of the Phecode in the study population and misclassification rates (estimated from the manually compiled exclusion criteria list) of Phecodes with greater than 5% absolute bias.

Further, to compare the performance of Logistic regression with exclusion criteria (Gold-standard model) and the proposed Poisson model, we conducted analyses with the four SNPs previously published in the PheWAS papers (Denny et al. [2]), rs660895 (Figure 5), rs1847134 (Supplementary Figure 2), rs258322 (Supplementary Figure 3), rs4977574 (Supplementary Figure 4). In each figure, we illustrate the results from Gold-standard logistic regression, bias-corrected Poisson model and Poisson model with robust sandwich standard errors (Stock and Watson [9]). We controlled the statistical significance at FDR = 0.1 level (p-values ≤ 1.20 x 10^(9^). Note that currently, the robust standard errors option is not compatible with the bias-correction option. In our previous simulation studies (not shown), the bias-correction procedure only affects codes with infinite estimates, which constitutes less than 0.5 percent of the codes in real data. Lastly, we implemented the proposed Poisson model on ICD 9 codes with the same SNPs used in PheWAS analyses. Age, gender and the first three genetic principal components were included as covariates for PheWAS and ICD 9 analyses. The results are shown in Figure 6, Supplementary Figure 5, Supplementary Figure 6 and Supplementary Figure 7. We controlled the statistical significance at FDR = 0.1 level (p-values ≤ 4.16 x 10^(:^).

**Figure 5:**
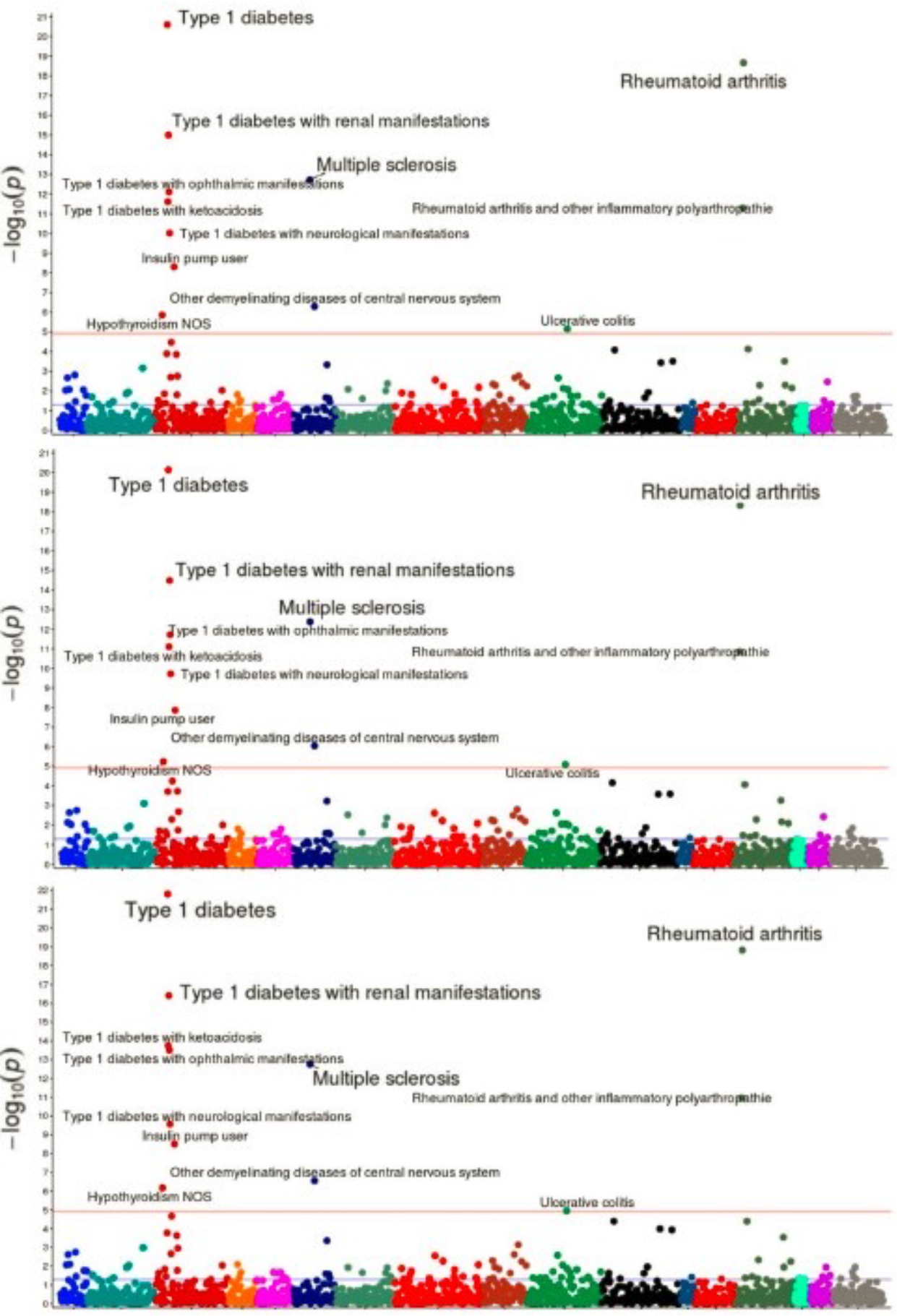
PheWAS analysis result for SNP rs660895 with Gold-standard Logistic regression model (top), bias-corrected Poisson (middle) and robust Poisson model (bottom). Each dot represents one Phecode. The red line indicates the FDR significance line. X axis denotes the negative log 10 p- values. Top selected codes are Type I diabetes, Rheumatoid arthiritis, Type I diabetes with renal manifestations and Multiple sclerosis. The maximum vaulue of X axis is 21 for the top two plots and 22 for the bottom plot.

**Figure 6:**
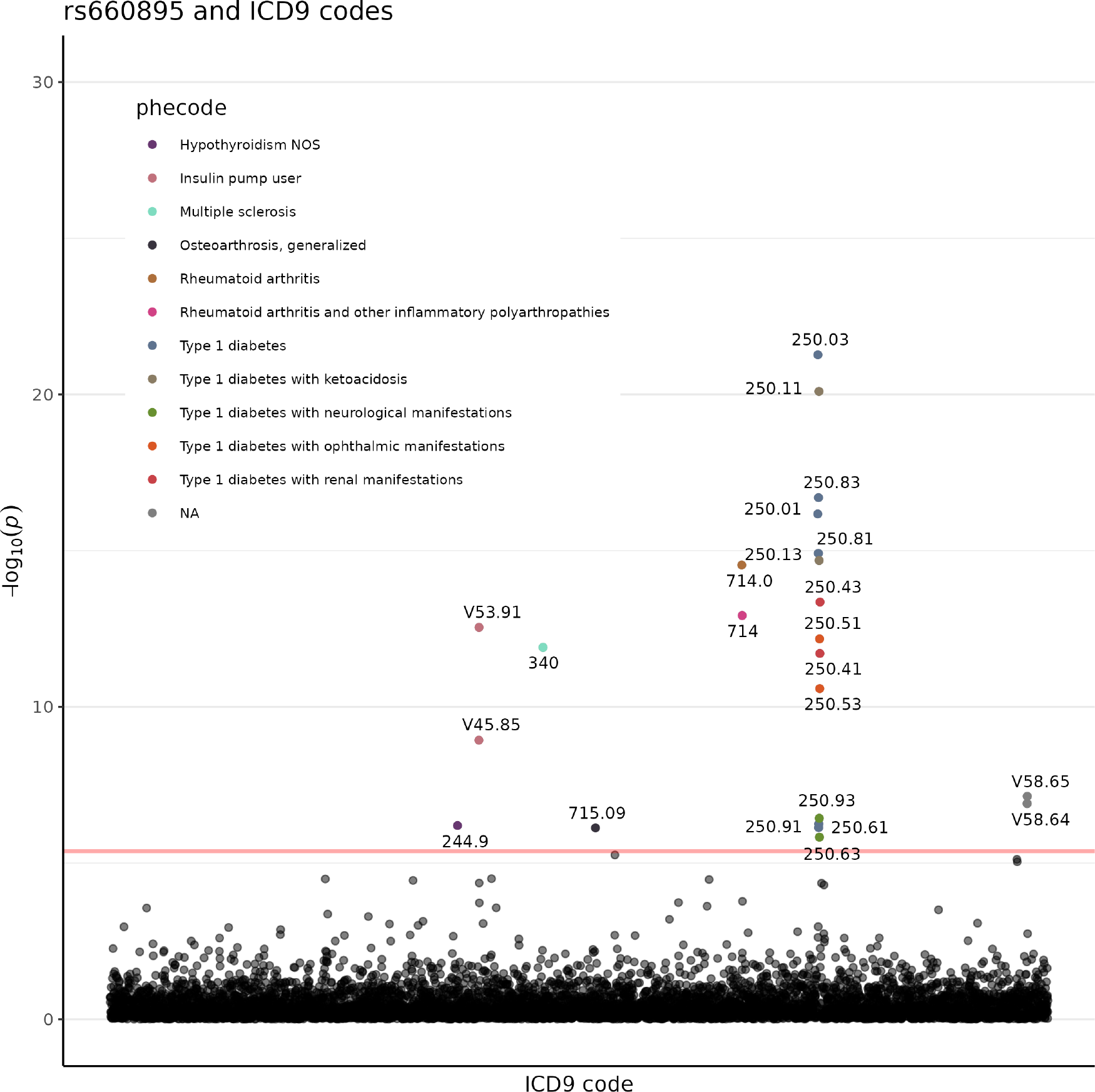
Result for ICD 9 analysis with SNP rs660895. Each dot represents an ICD 9 code. Y-axis denotes the negative log 10 P- values. The color of the dot corresponds to Phecode groups indicating in the legend. The red line indicates the FDR significance line. The group NA means that the codes do not correspond to any Phecode group. The significant ICD 9 codes in the red boxes correspond to V42.83, Pancreas replaced by transplant and V58.64, Long-term usage of non-steroidal anti-inflammatory. These two codes do not belong to any Phecode group.

### 4.3 Clinical Application Results

In the Simulation Results section, biases mainly occurred when the prevalence of the outcome was high. The biases increased as misclassification rates increased. In Figure 4, with SNP rs14483486 as an example, Phecodes with more than 5% biases include common diseases like diabetic and hypertension related diseases with prevalence > 0.2. In addition, the misclassification rates for these Phecodes are high, more than 0.85. The observations match the trend in the simulation results. Since we do not know the true classification of the control population, the prevalence and misclassification rates were calculated assuming the observations removed after exclusion criteria were true cases, misclassified as controls in the original data. Take Phecode 250.7 for example, there were 34592 subjects classified as true controls, 574 as true cases and 9598 observations removed. We assumed that subjects removed were true cases. The estimated prevalence was (574 + 9598)/44764 = 0.227. The estimated misclassification rate was 9598/ (9598+574) = 0.943.

To compare the performance of Poisson models and Gold-standard model, we conducted PheWAS analyses on the BioVU data with 4 SNPs. In general, Poisson models obtain comparable inference to Gold-standard model. Gold-standard model captures a large number of disease-genetic associations including Type I diabetes and Rheumatoid arthritis related Phecodes with SNP rs660895 (Figure 5, top). Both bias-uncorrected Poisson model with robust standard errors (Figure 5, bottom) and the bias-corrected Poisson model (Figure 5, middle) can obtain all the significant Phecodes captured in Gold-standard method. In Supplementary Figure 2, Gold-standard model only captures one Phecode, “Other non-epithelial cancer of skin” with SNP rs1847134 (Supplementary Figure 2, top). Both bias-uncorrected Poisson model with robust standard errors (Supplementary Figure 2, bottom) and bias-corrected Poisson model (Supplementary Figure 2, middle) can catch the Phecode. In Supplementary Figure 3, although both Poisson models fail to get the Phecode, “Neoplasm of uncertain behavior of skin”, captured by the Gold-standard model with SNP rs258322 (Supplementary Figure 3, top), they can obtain all other skin cancer related Phecodes (Supplementary Figure 3, middle and bottom). In Supplementary Figure 4, bias-uncorrected Poisson model with robust standard errors (Supplementary Figure 4, bottom) is able to catch all-important Phecodes captured in Gold-standard model about Coronary artery disease (Supplementary Figure 4, top). the Phecode, “Unstable angina”, close to the significance borderline, is missed by bias-corrected Poisson model (Supplementary Figure 4, middle).

Since the estimation of relative risk can bypass the need for exclusion criteria lists, we further extend Poisson model to large-scale phenomes. We conducted analyses on ICD 9 codes via bias-uncorrected Poisson model with robust standard errors. The corresponding ICD 9 codes analysis result for Supplementary Figure 2 is shown in Supplementary Figure 5. The ICD 9 codes obtained belong to non-epithelial skin cancer related Phecode groups, “Other non-epithelial cancer of skin” and “Neoplasm of uncertain behavior of skin”, similar to the result obtained in PheWAS analysis (Supplementary Figure 2). The corresponding ICD 9 codes analysis result for Supplementary Figure 3 is shown in Supplementary Figure 6. The ICD 9 codes obtained belong to epithelial skin cancer and skin cancer related to sun exposure Phecode groups, similar to the result obtained in PheWAS analysis (Supplementary Figure 3). The corresponding ICD 9 codes analysis result for Supplementary Figure 4 is shown in Supplementary Figure 7. The ICD 9 codes obtained belong to Coronary artery diseases Phecode groups, including “Coronary atherosclerosis”, “Angina pectoris” and “Unstable angina”, similar to the result obtained in PheWAS analysis (Supplementary Figure 4). These results show that with ICD 9 codes analyses, we are able to discern the major related diseases as in PheWAS analyses while obtaining more detailed description of the diseases. In addition, with ICD 9 codes analyses, we are able to capture information that might not be available in Phecode groupings. Take SNP rs660895 for example, the corresponding ICD 9 codes analysis result is shown in Figure 6. All but two ICD 9 codes obtained belong to the Phecode groups captured in PheWAS analysis (Figure 5). ICD 9 analysis captures 2 additional ICD 9 codes that were not previously defined in any Phecode group, denoting as “NA” (Figure 6).

These two ICD 9 codes are V42.83 (Pancreas replaced by transplant) and V58.64 (Long-term (current) use of non-steroidal anti-inflammatories). Based on the descriptions, these two ICD 9 codes relate to the surgical and drug-usage aspects of diabetes which are highly correlated with the Type I diabetes related Phecodes captured in Figure 5.

## 5 DISCUSSION

PheWAS analyses was invented to give a quick scan to assess disease-genetic relationship. Model that can handle binary outcomes is usually used, namely, the logistic regression model. In our setting, the outcomes are the aggregation of the billing codes in EMR data which are known to be noisy. To use the logistic regression model, strict assumption on the accuracy of the defined true “case” and “control” labels is required. It is our belief that more cares have been given to defining a case. The misclassification happens mainly in the control group. Efforts have been made in Denny et al. [2] to manually compile the exclusion criteria so that the correct estimates of odds ratio can be obtained. They were able to replicate several SNP-disease relationships as in the GWAS catalog. However, the manual compilation of the exclusion criteria has several drawbacks, including heavily time-consuming and being subjective. Although Phecodes are an aggregation of the ICD 9 codes, we lost some precision during the aggregation. The ICD 9 codes contain more information about a specific disease group and can provide more information for downstream analysis such as clustering. Nevertheless, it is generally not feasible to extend the manual compilation procedure to this number of codes or larger.

From the statistical standpoint, this issue can be viewed as misclassification in the control group for logistic regression. Different methods have been proposed to address this issue including validation data-based methods and model-based methods. In the validation data-based methods, the investigator could have part of the data being validated and obtained better sensitivity and specificity with these validated data (Edwards et al. [10]; Lyles et al. [11]). In PheWAS analyses, multiple outcomes are scanned. It is not easy to have validated data in the EMR system due to the large number of codes being compared. Further, if it takes similar time to obtain the validation dataset with compiling exclusion criteria, there’s no advantage of this method over the original Gold-standard method. The model-based methods incorporate the misclassification as a parameter in the models. By adjusting for the misclassification, the coefficients for the exposure should be unbiased. Liu and Zhang [12] proposed to incorporate misclassification parameter in the model and estimate the parameter with Fisher scoring algorithm. The simulation demonstrated that with the correct model specification, the odds ratio estimates are within 5% biases range. In addition, the method does not require any additional validation data. However, the algorithm doesn’t work well with higher misclassification rates with the potential of outputting invalid standard deviation estimates. In their simulation studies, the misclassification rate was only tested to 0.2. In PheWAS, the misclassification rate could range from 0.01 to 0.97. Therefore, more work needs to be done with the method to accommodate larger misclassification rate scenarios.

In this study, we evaluated the use of relative risk as an estimate that is appropriate in the EMR setting for binary outcomes. From the simulation, we observed that the biases occurred when there was misclassification in the control group without implementing any exclusion criteria. In addition, the biases increased as the prevalence of the outcome and misclassification increased. The relative risk estimates were unbiased in all the settings. The simulation result matched with the theoretical formula we derived. More, we confirmed in simulation that the unbiased relative risk can be either from a model that directly measures relative risk (Poisson model in our case) or conversion from the odds ratio, estimated from the logistic model. In the reality, we do not know the true estimates and misclassification rate for a code. It is likely that most of the codes have varying degrees of misclassification.

Without handling for the misclassification, the final estimates are altered. This can potentially influence the interpretation for the genetic-disease relationship in the later analysis.

When the relative risk model was used on the real data, the performance is similar to the Gold-standard method. Theoretically, when Poisson model is used on a binary outcome, it tends to be more conservative than the logistic regression. Indeed, there are other options with relative risk model, for example, the log-binomial model. Since it’s a transformation of the same distribution that was used in logistic regression, the variance should be correct.

However, it has been reported to behave poorly and fail to converge in some settings (Williamson, Eliasziw, and Fick [13]; Marschner and Gillett [14]). Fortunately, the over-conservative issue in the Poisson model can be overcome by applying the robust sandwich estimator (Stock and Watson [9]) to obtain the correct variance. This can be seen in SF8 where the Phecodes were all captured by Poisson with robust standard errors and less for the non-robust model. Another well-known issue for binary outcomes is the small cell count issue that leads to infinite estimate. The infinite estimates usually arise when one of the cells in a 2 × 2 table is 0. This can be corrected with implementing the Firth correction (Firth [8]). However, the Firth correction is not compatible with the robust standard errors. In all the SNPs and codes combination we analyzed, we rarely see codes with small cell counts issues. This could potentially be due to the pre-processing step following the Denny et al. [2] paper by excluding codes with less than 20 cases. If there are no infinite estimates, the bias-corrected relative risk is equivalent to the uncorrected one. To obtain better inference, it is usually recommended to apply robust standard errors with Poisson model if possible.

In the ICD 9 analysis, the ICD 9 codes captured by robust Poisson model match well with the Phecodes captured in the PheWAS analyses. The ICD 9 analysis is blinded with Phecode grouping information. This indicates that the model has enough power to obtain similar inference in ICD0 analysis as in the aggregated Phecodes analysis. In addition to capturing the important codes that match with their Phecode groups, more than 1 ICD 9 codes could be captured for 1 Phecode group. This indicates that we are able to learn what conditions of the disease contribute most to a certain disease group. Phecodes are aggregation of billing codes and not all ICD 9 codes are used to define Phecodes. Therefore, this information was lost in PheWAS analyses. In Figure 6, 2 codes are captured with no corresponding Phecodes group. However, these 2 codes contain information that are tightly related to other disease codes captured in the same analysis. Code V42.83 relates to Pancreas replaced by transplant. This seems to be a surgery code that relates to diabetes which is one of the main diseases captured in this SNP-disease analysis. Code V58.64 relates to the long-term usage of non-steroidal anti-inflammatory, which is a pain reliever that has been used mostly for patients with arthritis pain (Crofford [15]; S. Wongrakpanich et al. [16]). Several arthritis related codes are also captured in the same analysis. This information relate to the treatments and procedures the patients have undergone and could be very useful information for downstream analysis to assess the similarity between patients.

In this study, we evaluated relative risk as an alternative estimate to assess SNP-disease association that can overcome the misclassification issues in billing codes without manual compilation of any exclusion criteria list. Although several methods have been proposed to solve the misclassification issue in odds ratio estimation, additional issues still exist in the context of EMR data. Relative risk model provides unbiased estimates under current settings without considering the misclassification in the control group. The model is implemented in almost every existed software and ready to use. With this model, we are able to extend the PheWAS analyses to ICD 9 analysis without any restriction and able to obtain useful information that aren’t captured originally. This indicates that as more and more disease codes are categorized with more detailed information (i.e, ICD 10), relative risk as an estimate is able to be extended to analysis to larger database directly and have enough power to obtain the SNP-disease association.

## Data Availability

All data produced in the present study are available upon reasonable request to the authors.

## Author Declarations

All relevant ethical guidelines have been followed and any necessary IRB and/or ethics committee approvals have been obtained.

Yes

All necessary patient/participant consent has been obtained and the appropriate institutional forms have been archived.

Yes

---

**Supplementary Figure 1:**
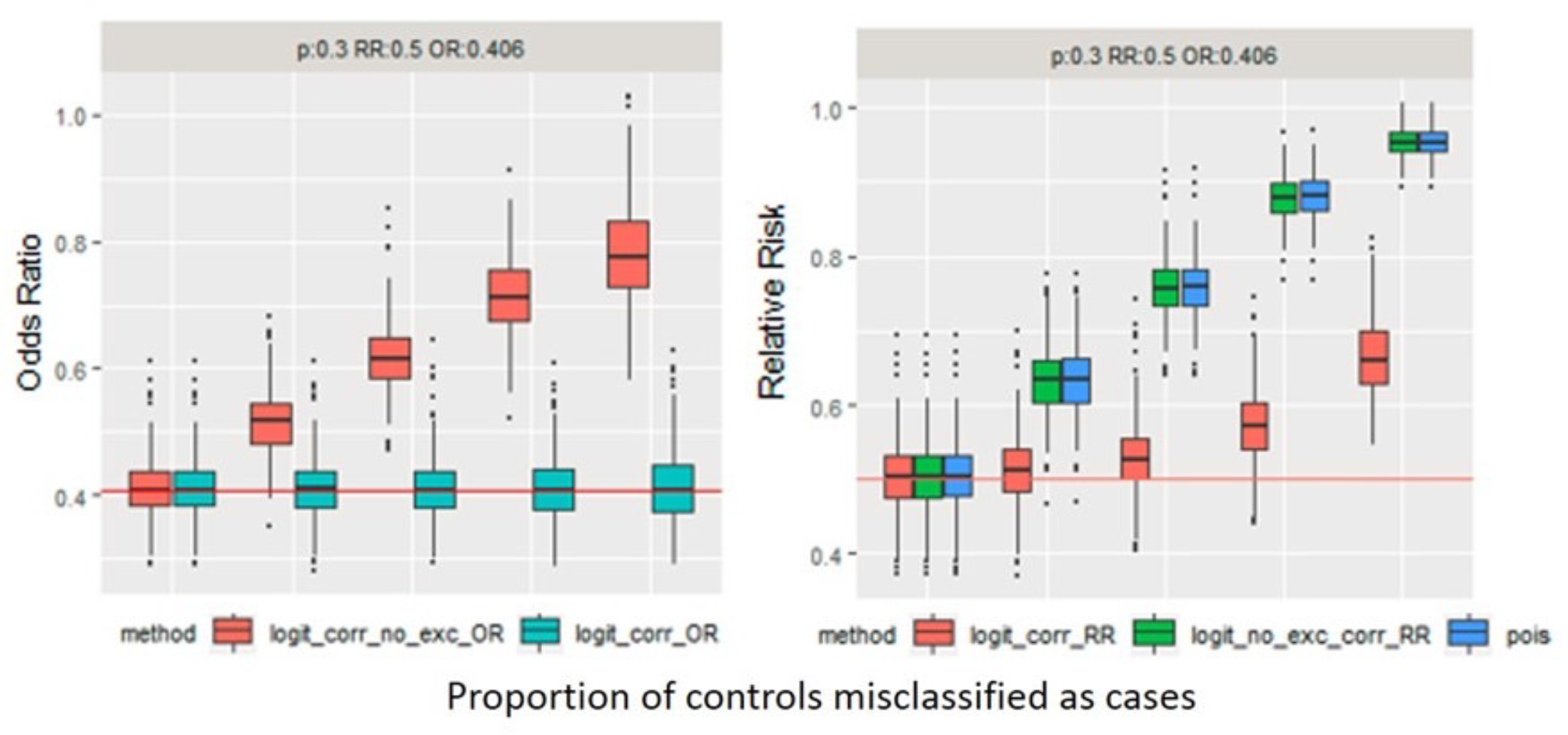
Univariate simulation result for controls misclassified as cases for odds ratios (left) and relative risk ratios (right). The prevalence = 0.3 and the exposure probability = 0.1. The true relative risk = 0.5 and the corresponding odds ratio = 0.406. The x-axis denotes the proportion of controls being misclassified as cases. the y-axis for the left plot denotes the odds ratios and relative risk estimates for the right plot. For the left plot, the red boxes correspond to odds ratios estimated from logistic regression without exclusion criteria. The teal boxes correspond to odds ratio estimated from logistic regression with exclusion criteria. For the right plot, the red boxes correspond to the converted relative risk estimates from logistic regression with exclusion criteria. The green boxes correspond to the converted odds ratio from the logistic regression without exclusion criteria. The blue boxes correspond to the relative risk estimated from Poisson model

**Supplementary Figure 2:**
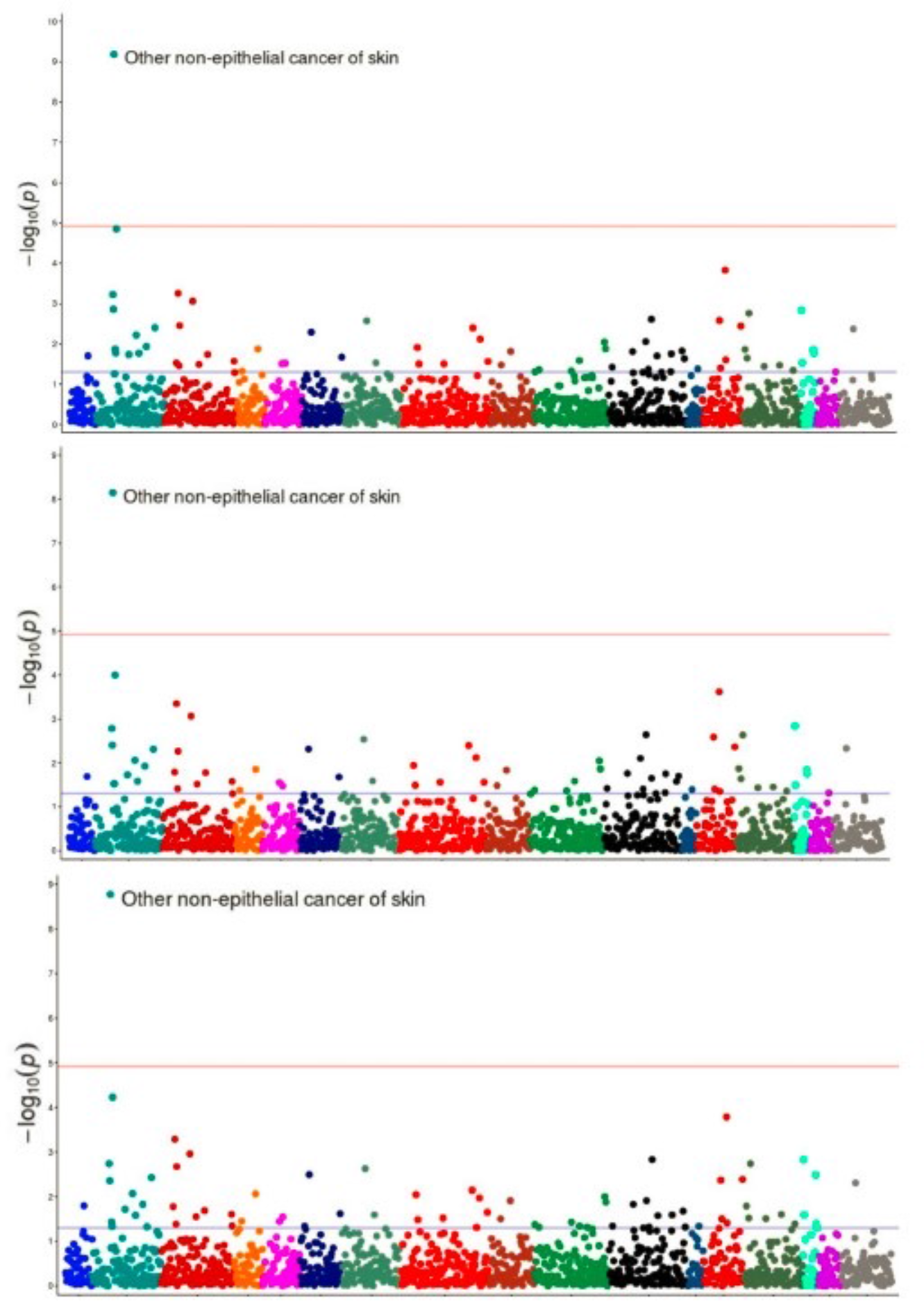
PheWAS analysis result for SNP rs1847134 with Gold-standard Logistic regression model (top), bias-corrected Poisson (middle) and robust Poisson model (bottom). Each dot represents one Phecode. The red line indicates the FDR significance line. X axis denotes the negative log 10 p-values. The only selected code is Other non-epithelial cancer of skin. The maximum vaulue of X axis is 10 for the top plot and 9 for the middle and bottom plot.

**Supplementary Figure 3:**
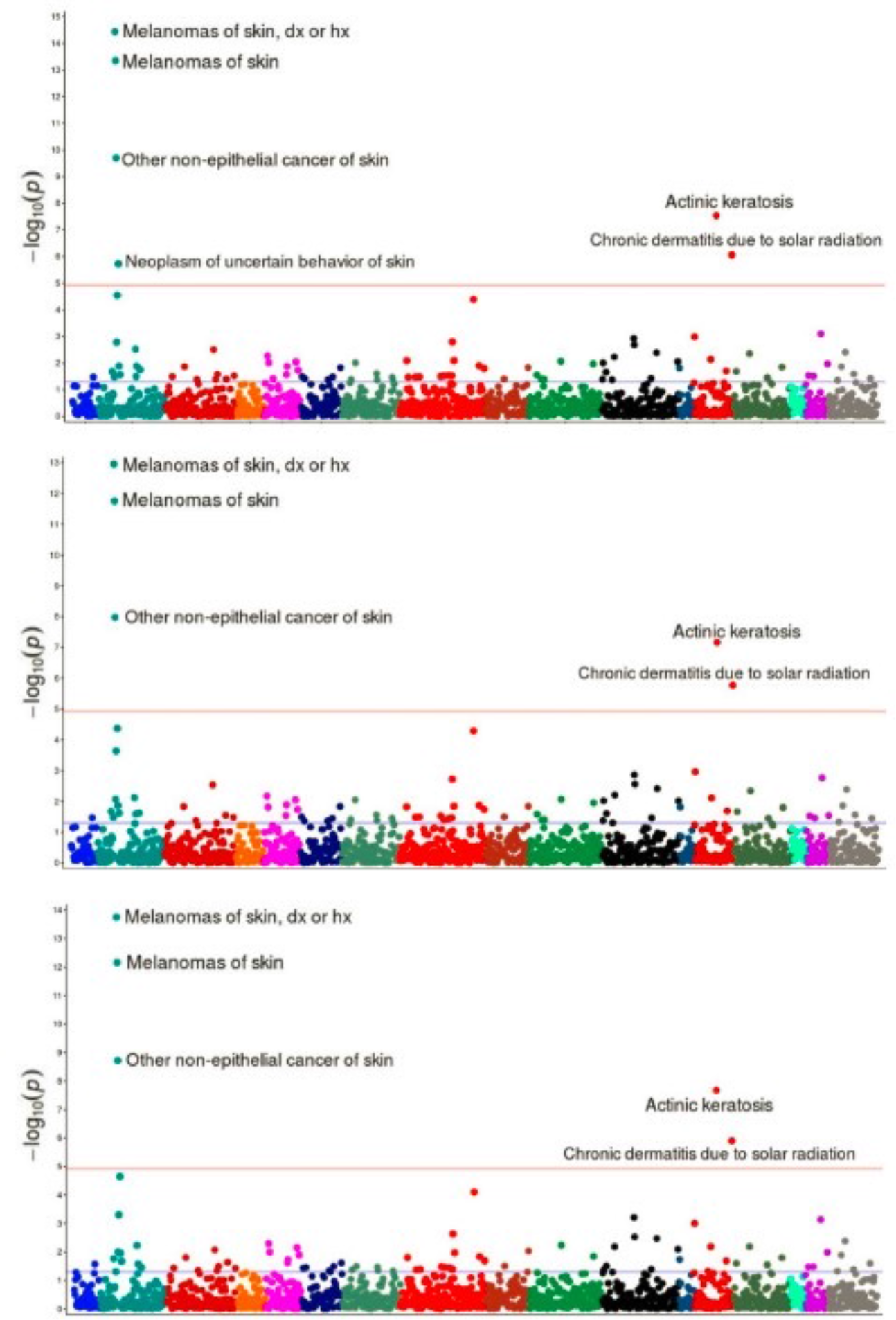
PheWAS analysis result for SNP rs258322 with Gold-standard Logistic regression model (top), bias-corrected Poisson (middle) and robust Poisson model (bottom). Each dot represents one Phecode. The red line indicates the FDR significance line. X axis denotes the negative log 10 p-values. Top selected codes are Melanomas of skin, dx or hx, Other non-epithelial cancer of skin and Actinic Keratosis. The maximum vaulue of X axis is 15 for the top plot; 13 for the middle and 14 for the bottom plot.

**Supplementary Figure 4:**
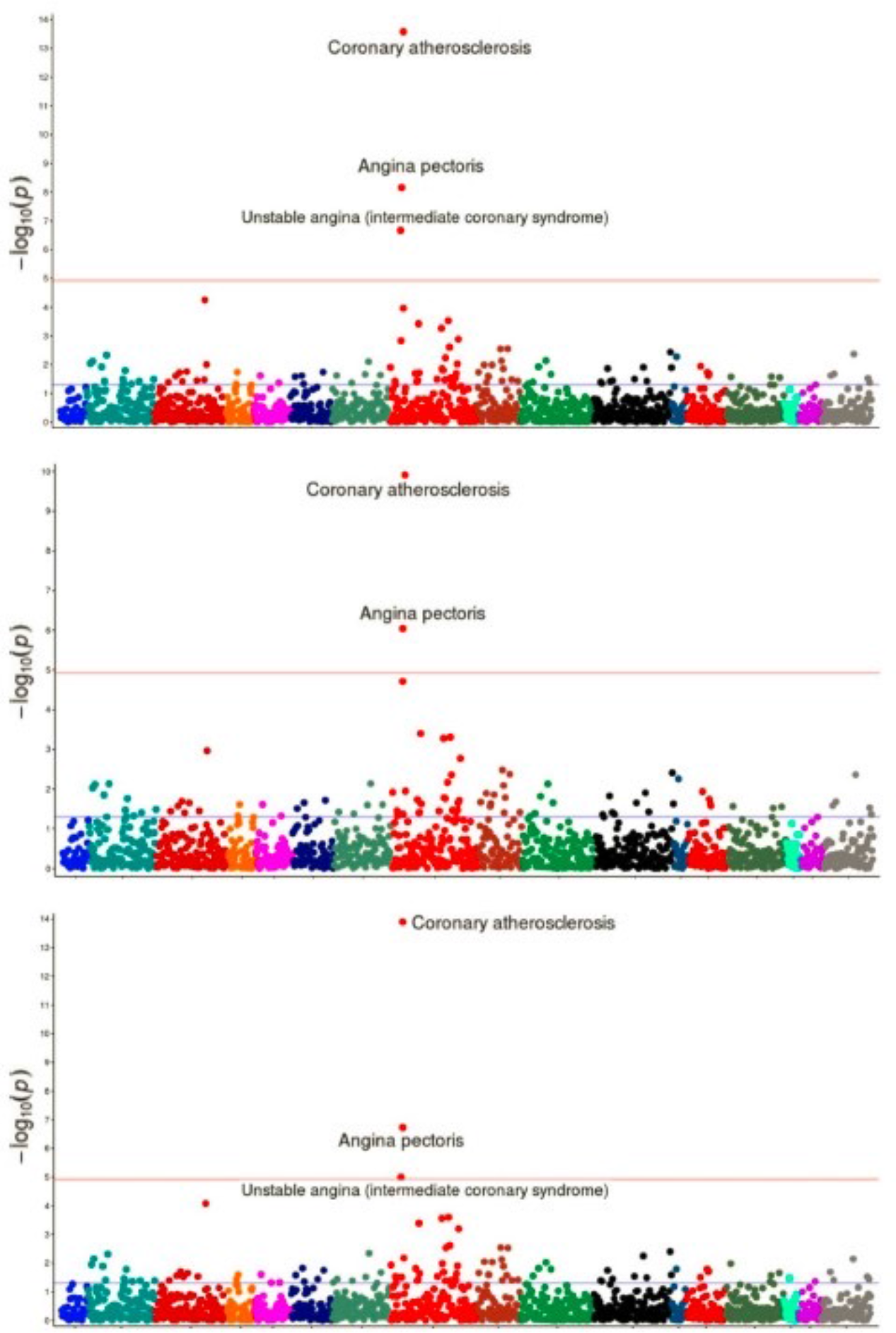
PheWAS analysis result for SNP rs4977574 with Gold-standard Logistic regression model (top), bias-corrected Poisson (middle) and robust Poisson model (bottom). Each dot represents one Phecode. The red line indicates the FDR significance line. X axis denotes the negative log 10 p-values. Top selected codes are Coronary atherosclerosis, Angina pectoris and Unstable angina. The maximum vaulue of X axis is 14 for the top plot; 10 for the middle and 14 for the bottom plot.

**Supplementary Figure 5:**
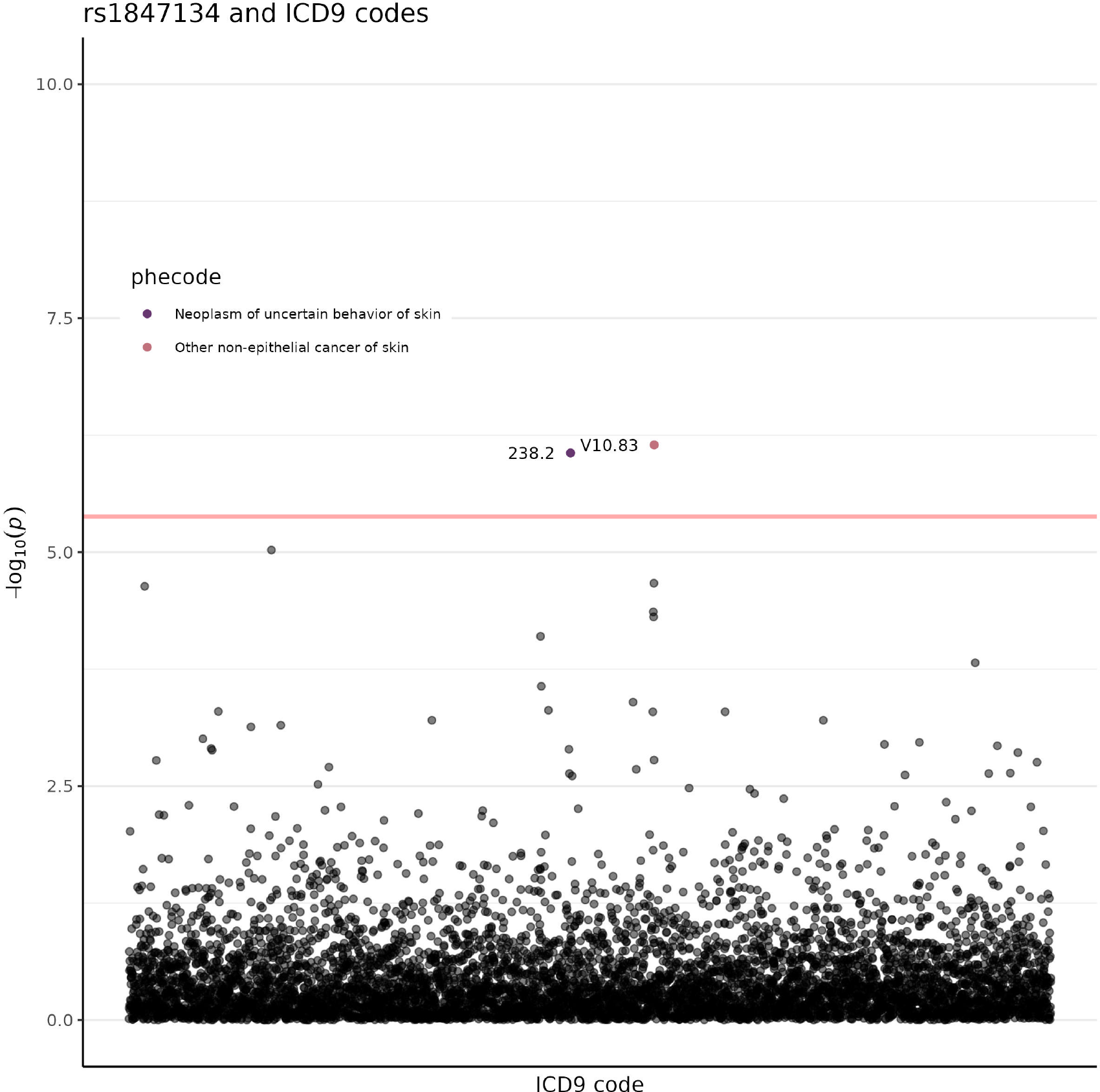
Result for ICD 9 analysis with SNP rs1847134. Each dot represents an ICD 9 code. Y-axis denotes the negative log 10 P-values. The colors of the dot corresponds to Phecode groups indicating in the legend. The red line indicates the FDR significance line.

**Supplementary Figure 6:**
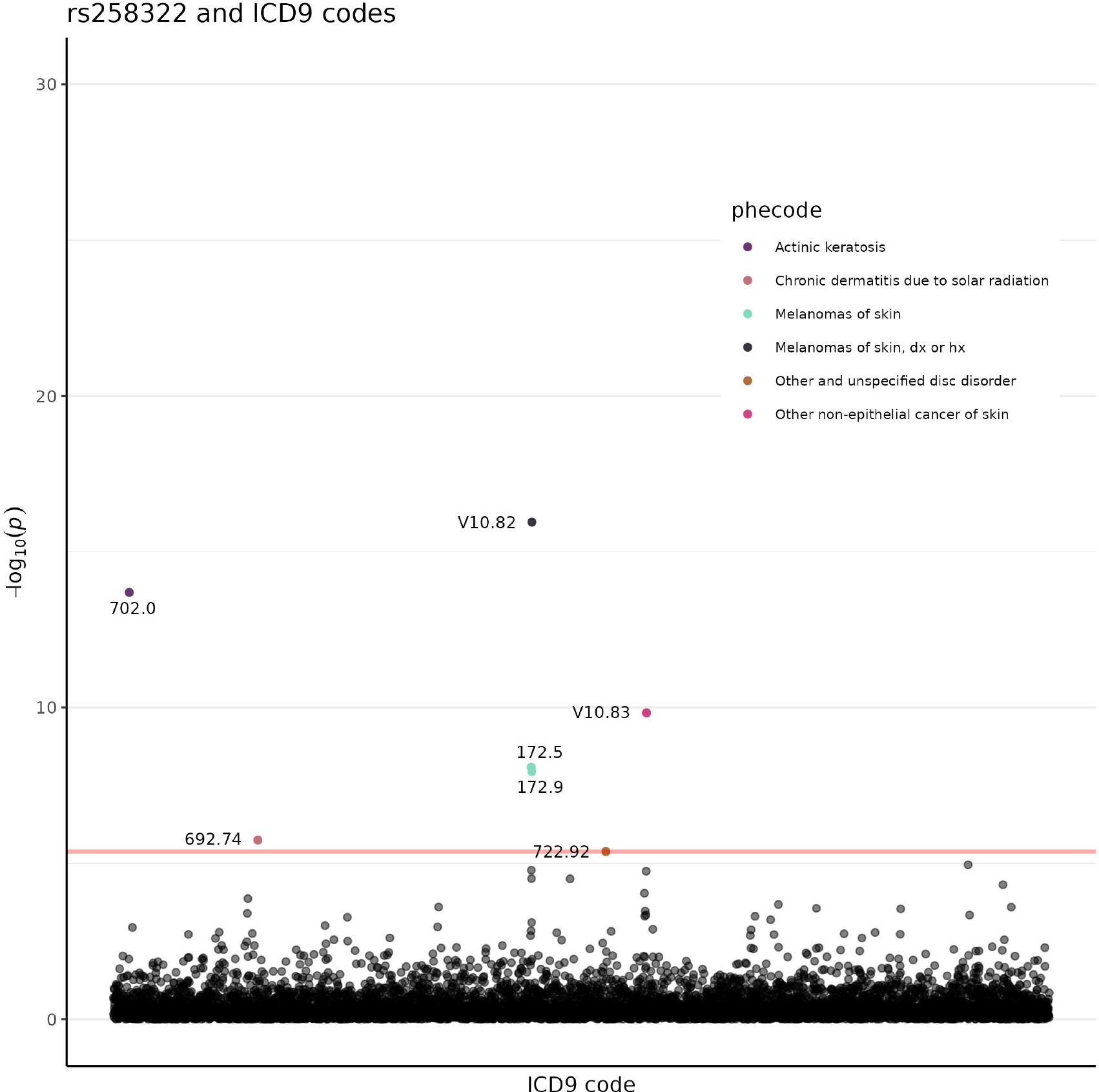
Result for ICD 9 analysis with SNP rs258322. Each dot represents an ICD 9 code. Y-axis denotes the negative log 10 P-values. The colors of the dot corresponds to Phecode groups indicating in the legend. The red line indicates the FDR significance line

**Supplementary Figure 7:**
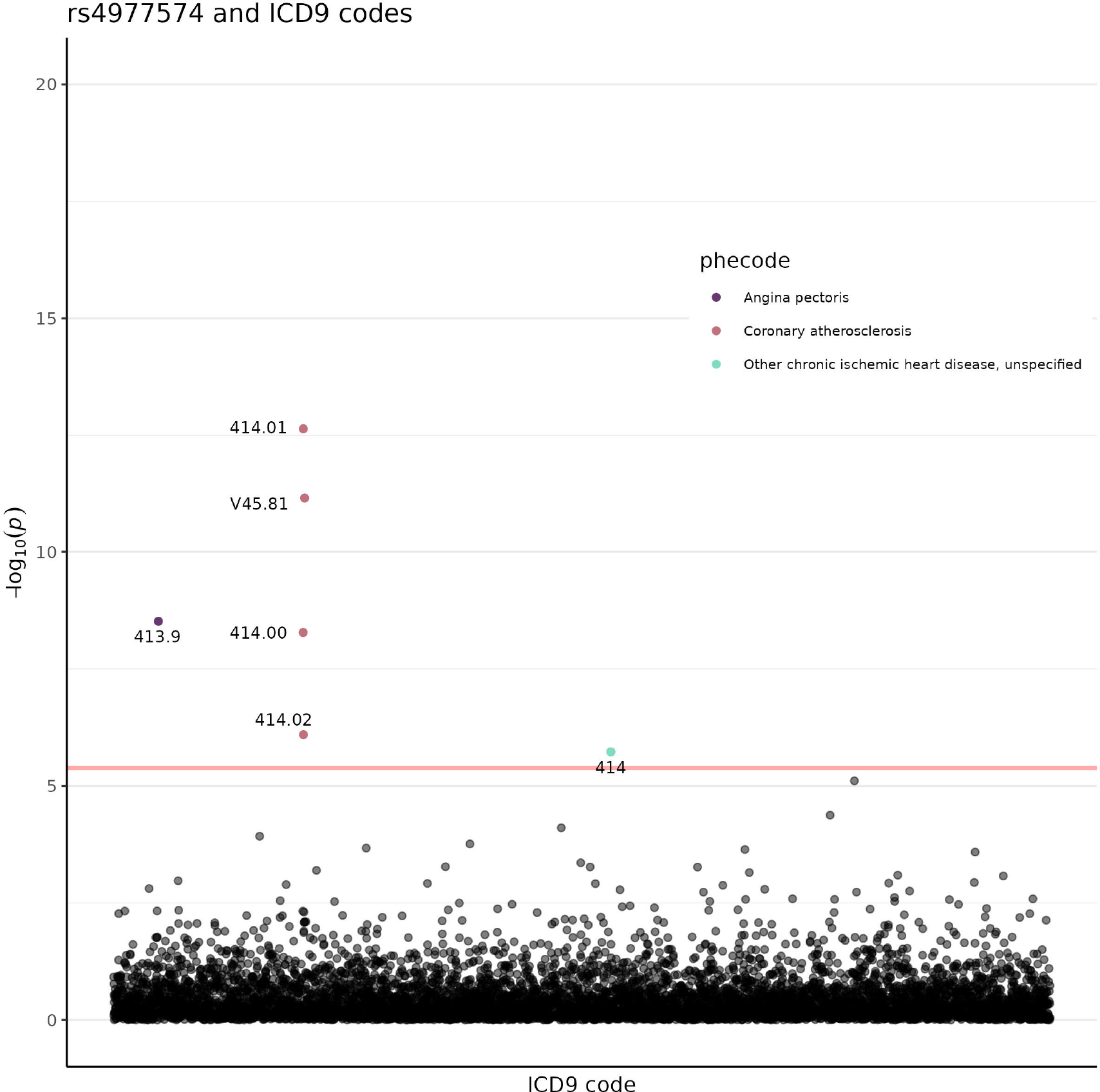
Result for ICD 9 analysis with SNP rs4977574. Each dot represents an ICD 9 code. Y-axis denotes the negative log 10 P-values. The colors of the dot corresponds to Phecode groups indicating in the legend. The red line indicates the FDR significance line.

